# The Genomic Architecture of Circulating Cytokine Levels Points to Drug Targets for Immune-Related Diseases

**DOI:** 10.1101/2024.04.19.24306036

**Authors:** Marek J. Konieczny, Murad Omarov, Rainer Malik, Tom G. Richardson, Sebastian-Edgar Baumeister, Jürgen Bernhagen, Martin Dichgans, Marios K. Georgakis

## Abstract

Circulating cytokines orchestrate immune reactions and are promising drug targets for immune-mediated and inflammatory diseases. Exploring the genetic architecture of circulating cytokine levels could yield key insights into causal mediators of human disease. Here, we performed genome-wide association studies (GWAS) for 40 circulating cytokines in meta-analyses of 74,783 individuals. We detected 359 significant associations between cytokine levels and variants in 169 independent loci, including 150 *trans*- and 19 *cis*-acting loci. Integration with transcriptomic data point to key regulatory mechanisms, such as the buffering function of ACKR1 acting as scavenger for multiple chemokines and the role of TRAFD1 in modulating the cytokine storm triggered by TNF signaling. Applying Mendelian randomization (MR), we detected a network of complex cytokine interconnections with TNF-b, VEGF, and IL-1ra exhibiting pleiotropic downstream effects on multiple cytokines. Drug target *cis*-MR paired with colocalization revealed G-CSF/CSF-3 and CXCL9/MIG as potential causal mediators of asthma and Crohn’s disease, respectively, but also a potentially protective role of TNF-b in multiple sclerosis. Our results provide an overview of the genetic architecture of circulating cytokines and could guide the development of targeted immunotherapies.

## Introduction

Chronic inflammation contributes to multiple human diseases, including allergic and autoimmune diseases, cardiometabolic diseases, and cancer. Inflammatory proteins like cytokines, chemokines and growth factors (hereafter collectively referred to as “cytokines”) orchestrate the immune response underlying inflammation.^1,2^ Circulating cytokines are readily accessible and therefore attractive targets for therapeutic modulation, as they represent soluble ligands that execute downstream mechanisms through binding to membrane receptors or other circulating agents.^3^ While several immunotherapies targeting circulating cytokines have been successfully introduced into the clinic, the lack of efficacy in other indications and the usually associated susceptibility to infection underscore the need for targeted approaches.^4,5^ Prioritizing specific downstream mediators is critical to minimize safety signals and ensure adherence to a life-long pharmacotherapy.^5,6^

Recent advances in human genetics have enabled an *in silico* prioritization of drug targets^7,8^, with approval rates more than two times higher than targets without genetic support.^9^ Mendelian randomization (MR) uses data from genome-wide association studies (GWAS) and offers a statistical framework for exploring associations between variants in genes encoding drug targets and disease traits.^10^ Previous MR analyses have illustrated the potential of integrating GWAS data for circulating proteins, including cytokines, with disease outcomes to discover novel drug targets.^11–16^ However, existing efforts have been largely restricted by the small sample sizes of GWAS studies for circulating cytokines. For example, the largest-to-date targeted GWAS which focused specifically on, circulating cytokines included up to 8,293 individuals and allowed the detection of 27 significant genomic loci for 41 cytokines.^17^

Novel proteomic platforms, such as the aptamer-based SOMAScan^®^ and the proximity extension assay Olink^®^, gain popularity in quantifying at scale large numbers of proteins including cytokines. Here, we performed cross-assay comparisons in the genetic architecture of 40 cytokines quantified with 3 approaches (multiplex bead-based immunoassay, aptamer-based assay, proximity extension assay) and pooled data in GWAS meta-analyses including up to 74,783 individuals. This effort allowed the detection of 359 significant associations between 169 independent genomic loci and one or more of the 40 cytokines offering novel insights into mechanisms regulating circulating cytokine levels. Applying MR, we establish a causal cytokine network including upstream mega-regulator cytokines that exert influence on a range of other cytokines. Finally, integrating these data with GWAS data for relevant disease endpoints, we provide genetic support for putative anti-inflammatory drug targets.

## Results

### Study cohorts and cross-assay reproducibility rate of significant genomic loci

We leveraged summary-level GWAS data for 40 circulating cytokines from 3 published datasets summing up to 74,783 individuals: the Cardiovascular Risk in Young Finns Study (YFS) and FINRISK studies that measured cytokines in serum using Luminex bead-based multiplex immunoassays (N=8,293); the Systematic and Combined AnaLysis of Olink Proteins (SCALLOP) study that measured cytokines in plasma using the proximity extension assay-based Olink^®^ platform (N=30,931); and the dataset provided by deCODE that measured cytokines in plasma using the aptamer-based SOMAScan^®^ assay (N=35,559, **Figure 1**).

**Figure 1.**
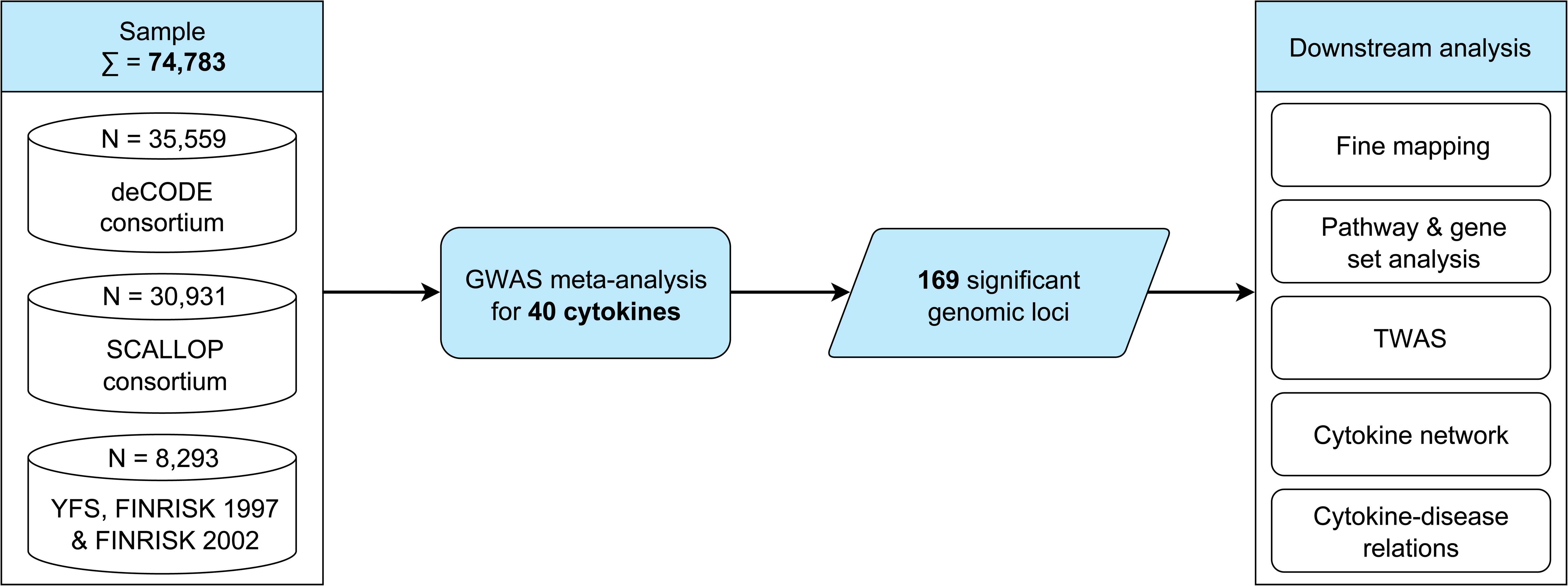
Flowchart of the study design. Illustration of the analytical pipeline steps applied in this study to decipher the genetic architecture of circulating cytokines and their relation to allergic and autoimmune, cardiometabolic and cancer outcomes. SCALLOP, Systematic and Combined AnaLysis of Olink Proteins; SNP, Single-nucleotide polymorphism; TWAS-MR, Transcriptome-wide Mendelian randomization analysis; YFS & FINRISK, Cardiovascular Risk in Young Finns Study.

Given the known differences across the assaying methods, we first tested the replication rate of significant variants detected in each dataset in the other two datasets.^18^ Although the GWAS in SCALLOP led to a considerably lower number of genome-wide significant loci for available cytokines, these variants showed the highest rate of reproducibility (p<5×10^-5^ and directionally consistent) in the other two datasets (median 67% in YFS & FINRISK and 63% in deCODE, **Figure 2**, **Supplementary Table S1**). A lower reproducibility rate was found for significant variants detected in YFS & FINRISK (median 4% in deCODE and 11% in SCALLOP) and deCODE (median 21% in YFS & FINRISK and 19% in SCALLOP). The cytokines that showed the highest relative proportion of reproducible SNPs across all 3 datasets independently of the measuring assay were CC chemokine ligand 2 (CCL2/MCP-1) and vascular endothelial growth factor (VEGF).

**Figure 2.**
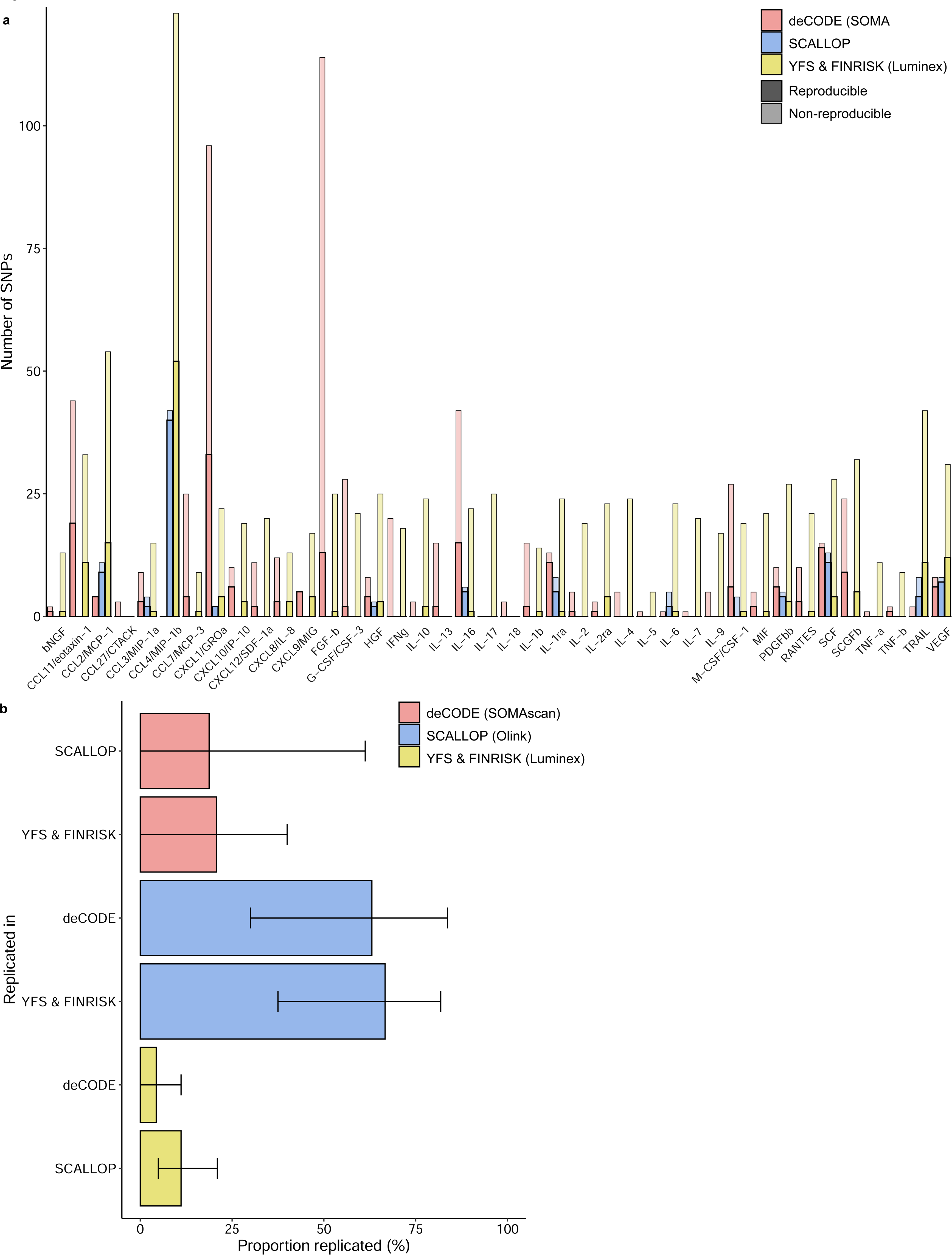
Comparisons of significant genomic loci for 40 circulating cytokines across 3 proteomics assays. **(a)** Number of reproducible and non-reproducible SNPs per cytokine (depicted as saturated and light-colored bars, respectively) for deCODE, SCALLOP and YFS & FINRISK cohorts. (**b**) Median proportion of replicated SNPs across the three platforms (error bars represent the 25^th^ and 75^th^ percentiles). Colored bars represent deCODE consortium in red, SCALLOP consortium in blue and YFS & FINRISK cohorts in yellow. SNP, Single-nucleotide polymorphism; SCALLOP, Systematic and Combined AnaLysis of Olink Proteins; YFS & FINRISK, Cardiovascular Risk in Young Finns Study.

### GWAS meta-analysis reveals novel *trans*-and *cis*-acting variants

Next, we performed GWAS meta-analyses across the 3 datasets. We identified a total of 359 significant associations between variants at 169 independent genomic loci and the circulating levels of one or more of the 40 cytokines (p<5×10^−8^ in fixed-effects meta-analysis, **Figure 3**, **Supplementary Table S2**). Variants that showed significant heterogeneity between the three cohorts (HetPval < 0.1) are reported in **Supplementary Table S3** (48% of the significant loci; range 0% to 100% across the 40 cytokines). The lambda values ranged between 0.96 for interleukin (IL)-16 and 1.04 for basic fibroblast growth factor (FGF-b), indicating absence of overall inflation in the test statistics (**Supplementary Table S3**). According to GWAS catalog (https://www.ebi.ac.uk/gwas/)^19^, 156 of the loci have not been associated with circulating levels of the 40 cytokines in previous GWASs (**Supplementary Table S4**). The proportion of explained variance by the significant variants ranged from 0.0008 for IL-17 to 0.033 for stem cell growth factor beta (SCGF-b) (**Supplementary Table S2**).

**Figure 3.**
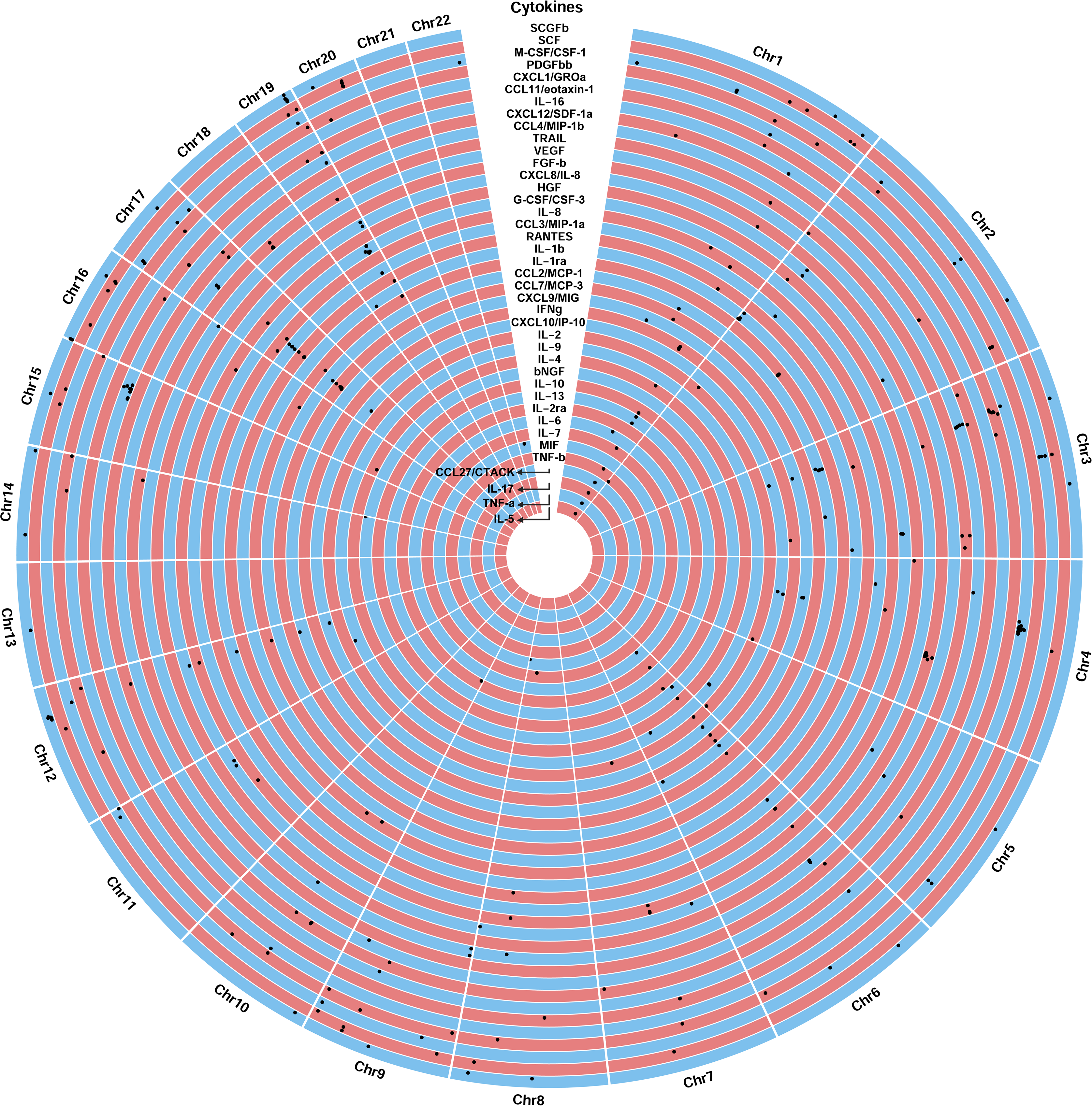
Genetic architecture of the circulating levels of the 40 cytokines. Circular Manhattan plot of genomic loci significantly associated with circulating levels of 40 cytokines in a meta-analysis of the three datasets. The 359 genome-wide significantly associated variants at p < 5×10^−8^ are depicted as black dots for GWAS meta-analyses in YFS & FINRISK, SCALLOP, and deCODE cohorts. The horizontal and vertical location of dots in each single rectangle depict genomic positioning (increasing from left to right) and p-value (descreasing from bottom to top), respectively. SCALLOP, Systematic and Combined AnaLysis of Olink Proteins; YFS & FINRISK, Cardiovascular Risk in Young Finns Study.

As expected due to larger effects sizes for rare genetic variants, we found a strong inverse correlation between minimum allele frequency and effect size (Spearman’s rho=-0.827, p=5×10^-30^, **Figure 4a)**. The majority of the significant loci (150 out of 169) represented *trans*-(distant-) acting variants. When excluding the human leukocyte antigen (HLA) region on chromosome 6, we found 33 pleiotropic variants showing associations with >1 cytokine among the significant *trans*-variants (**Figure 4b**). A locus hotspot associated with multiple cytokines was found at the region of the gene encoding *complement factor H* (*CFH*). This soluble mediator plays an essential role by interacting with the C3 convertase for regulation of inflammatory responses exerted by the complement system, which could possibly explain the associations with multiple cytokines.^20^ While at least 1 significant *trans*-variant was present for all studied cytokines (median number of variants per cytokine = 5, range 1 to 22), we found significant *cis*-(local-) acting variants in the vicinity of their encoding gene for 19 cytokines (**Supplementary Table S2**). The lead *cis*-acting variants showed stronger associations with cytokine levels (mean absolute beta: 0.18, range: 0.05-0.94) than *trans*-acting variants (mean absolute beta: 0.08, range: 0.03-0.55, *p*-for-comparison=0.03, **Figure 4c**).

**Figure 4.**
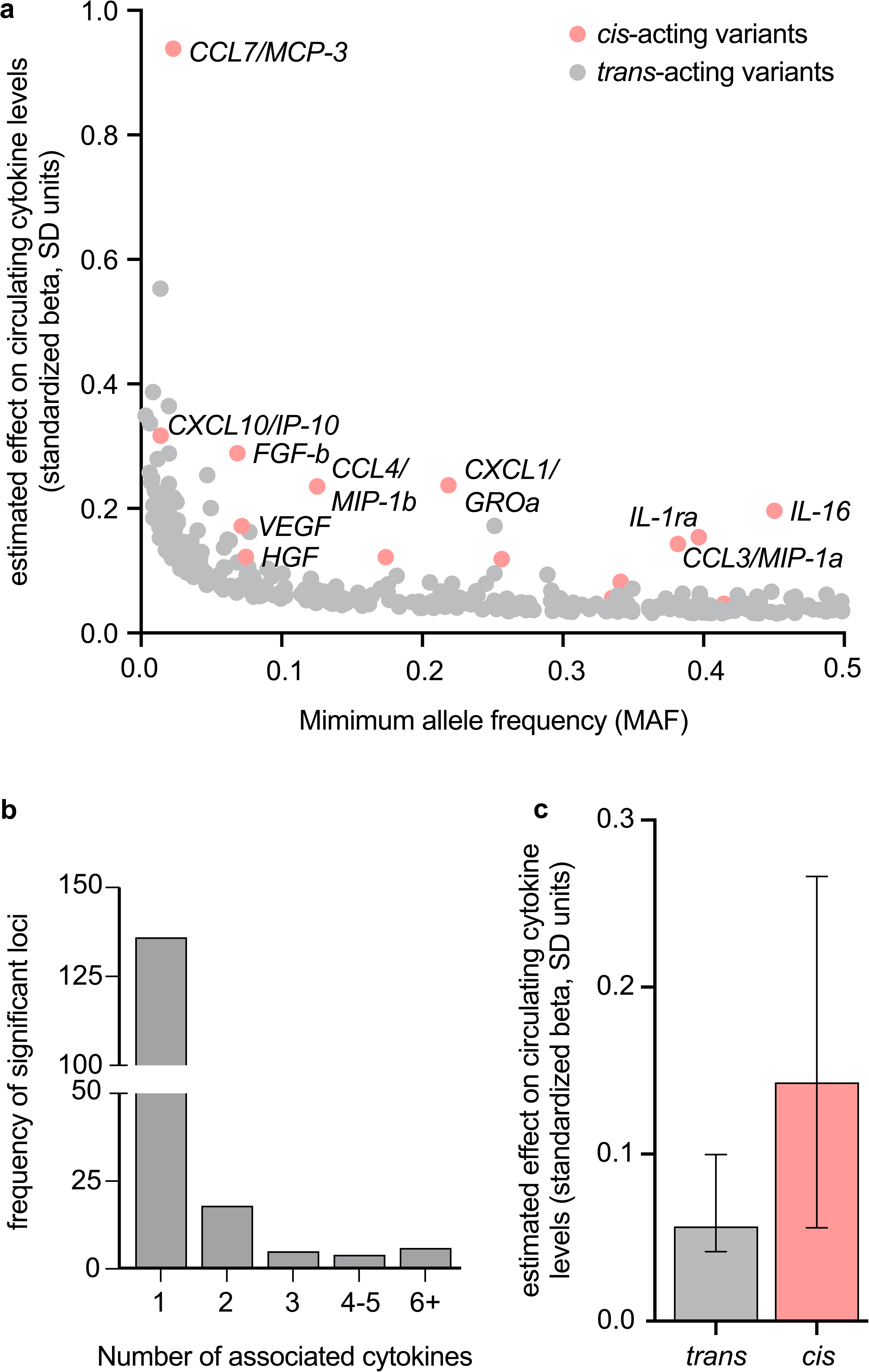
*Trans*-and *cis*-acting genetic variants underlying circulating cytokines. (**a**) Inverse correlation between minimum allele frequency and effect size illustrated for *trans*-and *cis*-acting loci. (**b**) Number of pleiotropic loci associated with circulating cytokines (excluding the HLA region on chromosome 6). (**c**) *Cis*-acting variants showed stronger associations with cytokine levels when compared with *trans*-acting variants. Bars and lines represent median and 95% confidence intervals, respectively. HLA, human leukocyte antigen; SD, standard deviation.

To map causal variants responsible for associations between cytokine serum levels and genes within each of the 169 independent genomic loci we used SuSiE fine mapping. Employing a Bayesian framework, fine mapping identifies credible sets of variants with posterior probability (PP) 95%. The number of variants within credible sets ranged from 2 to 50. The highest numbers of variants within a credible set were found at 15q21.3 for stem cell factor (SCF) (n=50), at 19q13.33 for SCGF-b (n=49), and at 6p21.1 for VEGF (n=44). SuSiE mapped the association test lead variant to the credible sets for 49 genomic loci, identifying the GWAS lead as most likely causal mutations **(supplementary Table S5).**

### Functional follow-up analyses highlight immune response regulatory mechanisms

To understand the biological significance and downstream functional impact of the identified variants, we performed follow-up analyses. A MAGMA gene-based analysis showed 829 significant associations with the levels of circulating cytokines at a Bonferroni-defined significance level (**Supplementary Table S6)**. In total, 626 uniquely mapped genes were associated with at least 1 cytokine. The number of genes mapped to cytokines ranged from 1 for beta nerve growth factor (bNGF), cutaneous T-cell attracting (CCL27/CTACK), IL-10 and tumor necrosis factor-alpha (TNF-a) to up to 95 genes mapped for SCGF-b, 92 genes for macrophage inflammatory protein-1β (CCL4/MIP—1b) and 51 genes for CCL11/eotaxin-1. In line with our GWAS results, the gene that was mapped for most cytokines (n=16) was *CFH*. The genes with the lowest p-value were *H4C14* for monocyte specific chemokine 3 (CCL7/MCP-3) (p=1×10^−50^), *DBA4* for IL-16 (p=1.3×10^−45^), and *ABC1* for SCF (p=1.9×10^−29^). A gene-property analysis revealed that the cytokine-related genes were primarily enriched for expression in the liver (p=4.9×10^−10^), in line with its well-established role as a main source of production of many cytokines. Other enriched tissues included the spleen (p=4.9×10^−4^) and lung (p=5.9×10^−4^, **Supplementary Table S7**). A MAGMA gene-set analysis prioritized 41 pathways for 12 cytokines that reached a Bonferroni-adjusted significance level (p<1.2×10^-7^, **Supplementary Table S8**). The identified pathways were primarily related to immune response with a small cluster involved in metabolic and developmental processes.

Positional mapping ascribed 75% of significant variants to intronic (54%) and intergenic (21%) regions, suggesting that the identified variants primarily determine gene transcription or gene expression profiles (**Supplementary Table S2**).^21^ Thus, we integrated our GWAS data with transcriptomic data and performed a transcriptome-wide association study (TWAS) using Mendelian randomization (MR) for a deeper elaboration on the transcriptional effects underlying our GWAS results. Summary statistics for expression quantitive trait loci (eQTLs) in whole blood were obtained from the eQTLGen consortium including transcriptomic profiles for 31,684 individuals of primarily European ancestry.^22^ Using *cis*-eQTLs as genetic instruments, we identified 245 significant associations between genetically proxied gene expression in whole blood and cytokines levels (**Figure 5**, **Supplementary Table S9**). The number of significant genes per cytokine ranged from 1 to 18. While most significant genes (78%) influenced the levels of a single cytokine, the genetically proxied expression of 54 genes showed an effect on circulating levels of up to 9 cytokines (n[*SKIV2L*]=9, n[5*HLA-DRB5*]=9, n[*NELFE*]=7, n[*ACKR1*]=5, n[*FCER1A*]=4, n[*TRAFD1*]=4, n[*LCMT2*]=4). Interestingly, we found significant *cis*-effects of the encoding gen expressions on the circulting levels of only 3 of the 40 respective cytokines. This is in line with previous eQTL-pQTL comparisons and aligns with the fact that the circulating proteome is not the direct product of the whole-blood transcriptome.^23,24^

**Figure 5.**
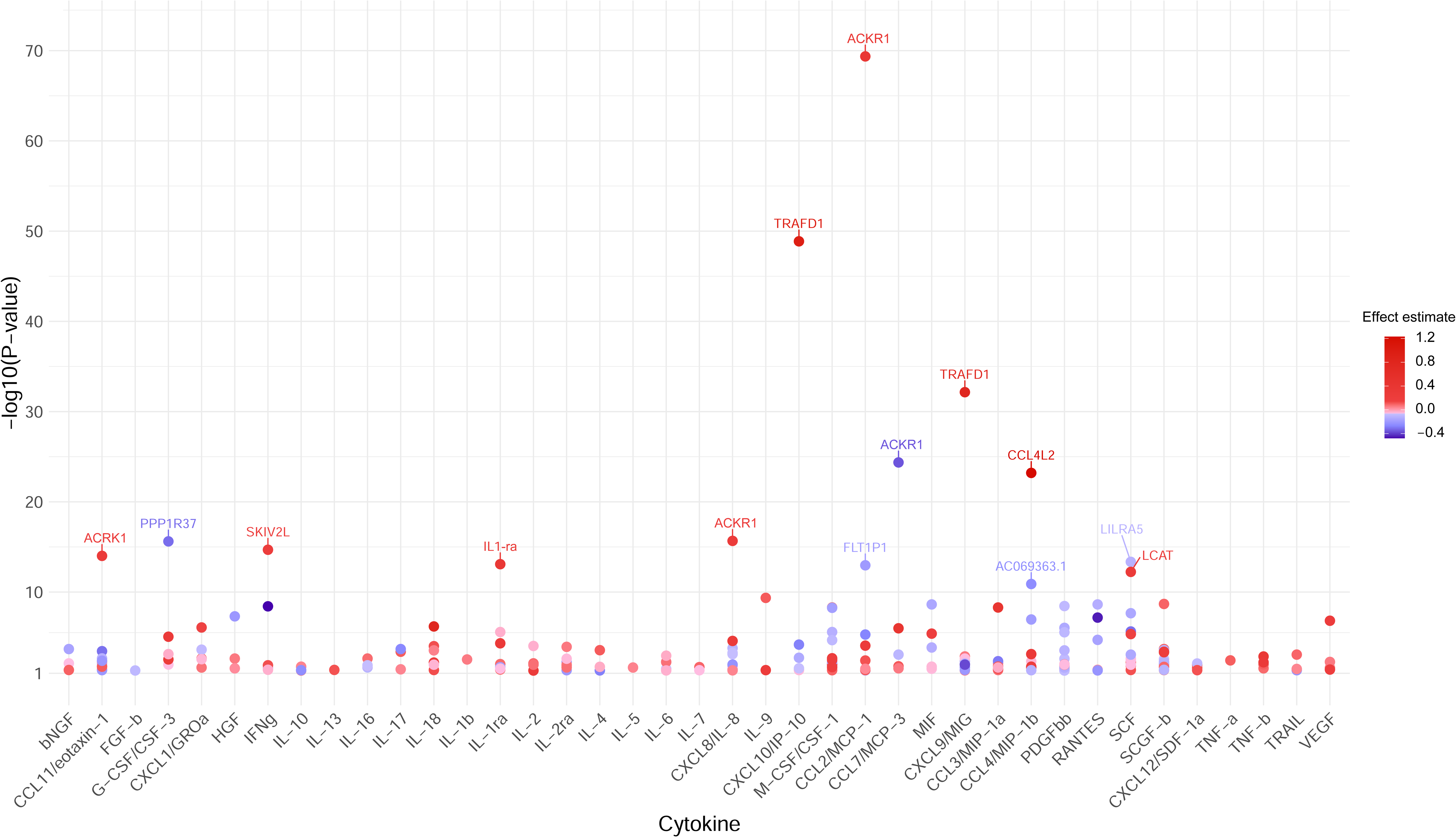
Genetically predicted gene expression in peripheral blood partly explains the genetic architecture of 40 circulating cytokine levels. The dots represent genes, the blood expression of which was significantly associated with circulating cytokine levels in a Mendelian randomization-based transcriptome-wide association study.

Excluding genes within the very dense HLA region (i.e. *SKIV2L, HLA-DRB5* and *NELFE*) we explored deeper the biological relevance of the pleiotropic *ACKR1*, *TRAFD1* and *LCMT2* genes. The genetically proxied mRNA levels of *ACKR1* were associated with circulating CCL2/MCP-1, CCL7/MCP-3, CCL11/eotaxin-1, growth regulated oncogene-α (CXCL1/GROa), CXCL8/IL-8. *ACKR1* codes for a cell-surface receptor that binds, internalizes and transports multiple CC and CXC chemokines and promotes leukocyte transcytosis into the circulation.^25,26^ By acting as scavenger receptor, ACKR1 modulates the bioavailability of cytokines and thereby affects inflammatory responses.^27,28^ The identified associations were driven by rs12075, a well-characterized missense variant in *ACKR1,* resulting in less efficient chemokine binding to ACKR1 due to loss of a necessary amino-acid sulfation (**Figure 6a**).^29^ The impaired receptor binding leads to elevated circulatory levels of chemokines and might in turn result in a compensatory increase in *ACKR1* expression which could explain the positive association between genetically proxied *ACKR1* and its ligands.^30^ We replicated previously reported associations between *ACKR1* and levels of CCL2/MCP-1, CCL7/MCP-3, CCL11/eotaxin-1, and CXCL1/GROa and additionally showed an association with CXCL8/IL-8 levels.^17,23^ The genetically proxied expression of *tumor necrosis factor receptor-associated factor 1* (*TRAFD1*) was also associated with multiple circulatory cytokine levels, including CCL7/MCP-3, monokine induced by interferon-gamma (CXCL9/MIG), interferon gamma-induced protein 10 (CXCL10/IP-10), and tumor necrosis factor-beta (TNF-b) (**Figure 6b**). TRAFD1 functions as an adaptor protein that binds to the intracellular domain of TNF receptors expressed on both innate and adaptive immune cells. It regulates downstream signaling also involving the NF-κB pathway and thereby modulates the production of several pro-inflammatory cytokines and inflammatory responses.^31–33^ TRAFD1 is a master regulator of genes involved in interferon-γ (IFNg) signaling and T-cell receptor activation.^34^ Genetically proxied expression of the gene encoding for LCMT2 showed associations with beta nerve growth factor (bNGF), CXCL8/IL-8, CXCL10/IP-10 and platelet-derived growth factor-bb (PDGFbb). LCMT2 is involved in amino-acid metabolism presumably regulating hypothalamic gene expression but there is only limited knowledge on its biological function.^35,36^

**Figure 6.**
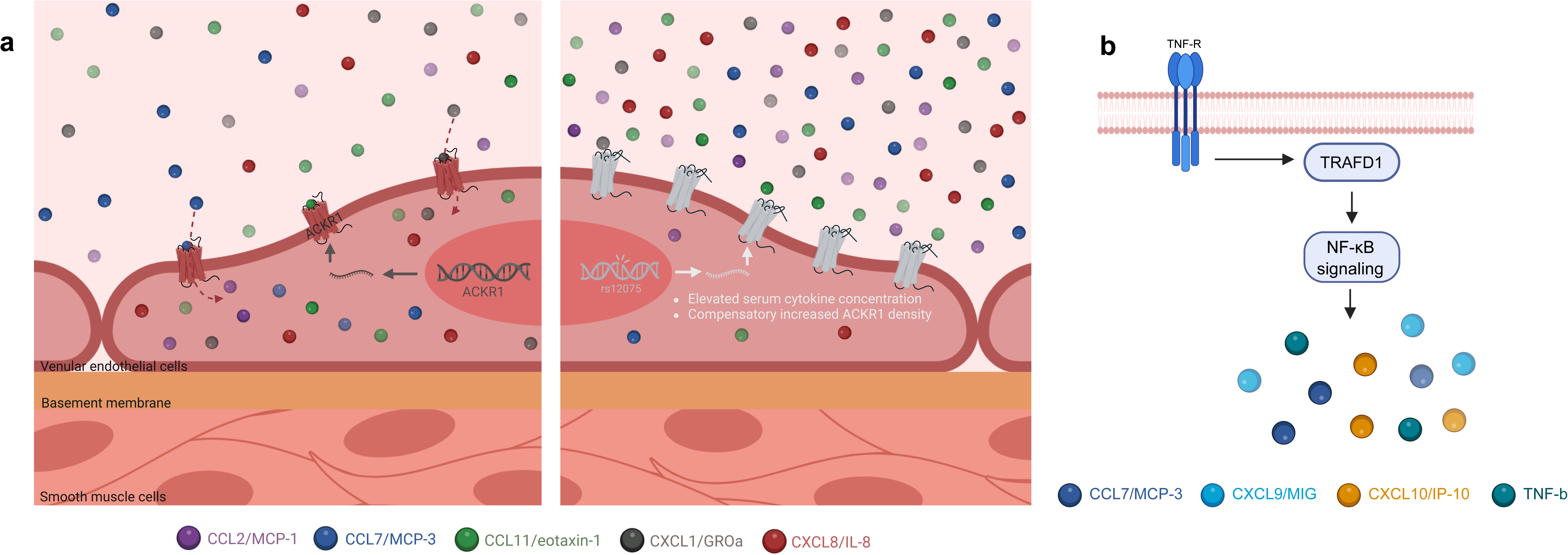
Gene expression of *ACKR1* and *TRAFD1* exert pleiotropic effects on multiple cytokine levels. (**a**) Schematic illustrating the impact of *cis*-eQTLs for *ACKR1* on receptor function. Left hand side shows *ACKR1* gene encoding the atypical chemokine receptor 1 functioning as sink for multiple chemokines which are buffered intracellularly in venular endothelial cells. Depicted on the right, is the missense variant rs12075 coding for a dysfunctional receptor with less efficient chemokine binding efficacy. This leads to higher levels of circulating CCL2/MCP-1, CCL7/MCP-3, CCL11/eotaxin-1, CXCL1/GROa, and CXCL8/IL-8 and possibly to a compensatory increase in *ACKR1* expression and receptor density. (**b**) Schematic illustrating how genetically proxied *TRAFD1* expression regulates multiple cytokine levels (CCL7/MCP-3, CXCL9/MIG, CXCL10/IP-10, TNF-b), supporting its regulatory role in TNF-mediated NF-κΒ signaling. ACKR1, atypical chemokine receptor 1; TNF-R, TNF-receptor.

### Genetic associations point to network interactions between circulating cytokines

As a next step, we explored cross-trait genetic correlations between the circulating levels of the 40 studied cytokines (**Figure 7a**, **Supplementary Table S10**). One third of the between-cytokine correlations were significant at p<0.05; the vast majority of the significant associations (96%) were positive. Furthermore, to understand causal interconnections between circulating cytokines levels, we performed MR analysis using *cis*-variants from our GWAS meta-analysis. We found significant (FDR-corrected p<0.05) associations between 65 cytokine pairs (53 positive associations and 12 negative associations, **Figure 7b**, **Supplementary Table S11**). Genetically proxied levels of CCL7/MCP-3, stromal cell-derived factor-1alpha (CXCL12/SDF-1a), granulocyte colony-stimulating factor (G-CSF/CSF-3), IL-9, TNF-b, and VEGF were positively associated with the levels of >2 other cytokines, whereas genetically proxied CXCL1/GROa and IL-1 receptor antagonist (IL-1ra) were negatively associated with lower level of >2 other cytokines. Most significant associations were detected for TNF-b (n=13), VEGF (n=9), IL-1ra (n=7), IL-9 (n=7), and G-CSF/CSF-3 (n=7). The negative associations between IL-1ra and several proinflammatory cytokines (CCL7/MCP-3, IL-9, TNF-a, TNF-b), chemokines (macrophage inflammatory protein-1α, CCL3/MIP-1a), and growth factors (hepatocyte growth factor, HGF; VEGF) align well with the immunoregulatory role of the IL-1 pathway and the inhibitory effect of IL-1ra on downstream IL-1 signaling.^37,38^. TNF-b emerged as a significant player in our network analysis, demonstrating characteristics of a master regulator by showing significant associations with higher circulating levels of 13 mostly pro-inflammatory cytokines. Furthermore, TNF-b exhibited significant positive LDSC genetic correlations with 7 of the 13 cytokines, suggesting a shared genetic architecture within the TNF-β network (**Figure 7a**). While certain interactions with TNF-b, such as those involving IL-1ra, TNF-a, TNF-related apoptosis inducing ligand (TRAIL), and VEGF, are well-documented, the majority of interactions have not been reported previously and merit additional investigation.^39–41^

**Figure 7.**
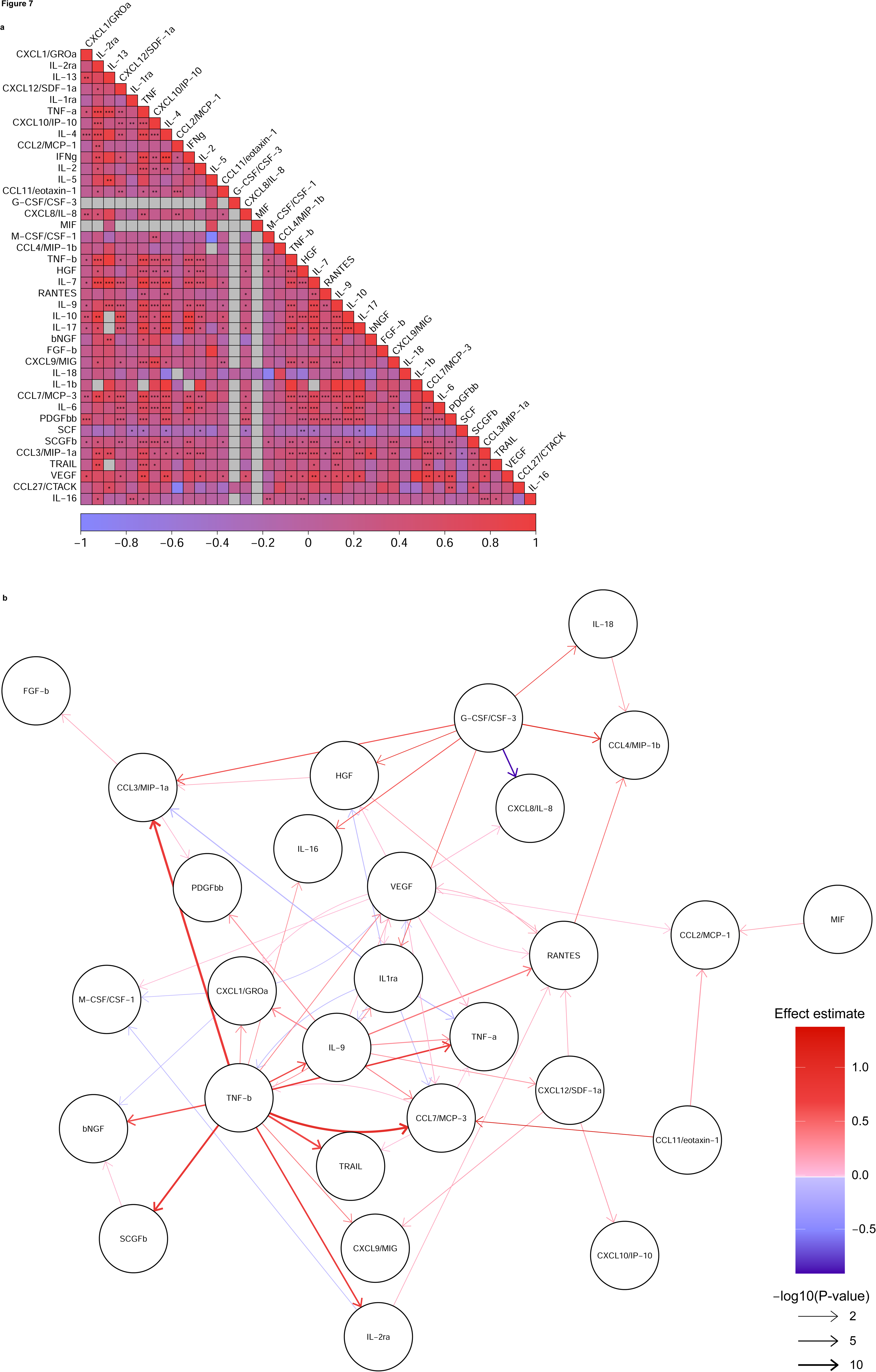
Cross-cytokine genetic associations. (**a**) Genetic correlations with LD-score regression across cytokine serum levels are depicted as correlation heatmap. Stars highlight significance level *, 0.05; **, 0.0001; ***, 0.00001. LD-score correlation coefficients are illustrated according to the legend below spanning from −1 in blue to +1 in red, missing correlation coefficients are depicted in grey. (**b**) *Cis*-Mendelian randomization between genetically proxied circulating cytokine levels. Arrow heads show the direction of causal influence, color gradient indicates the effect estimate and line width the logarithm-adjusted Benjamin-Hochberg corrected significance level. LD, linkage disequilibrium.

### *Cis*-Mendelian randomization and colocalization highlight potential drug targets for immune-related diseases

For insights into the clinical consequences of genetically proxied levels of the circulating cytokines, we analyzed associations with allergic and autoimmune, cardiometabolic, and cancer outcomes in two-sample MR followed by colocalization analyses (**Figure 8** and **Supplementary Table S12**). We used *cis*-acting genetic variants as instruments due to their lower likelihood of influencing cytokine levels through pleiotropic mechanisms. We further complemented these analyses with Bayesian colocalization to prioritize associations less likely to be influenced by pleiotropy due to linkage disequilibrium of studied variants with neighboring genes.^42^ Following correction for multiple comparisons, we found 24 significant MR associations between genetically proxied cytokine levels and disease outcomes (14 positive and 10 negative associations). Our MR findings partially confirmed established pathogenetic associations with diseases and therapeutic drug targets that are already in clinical application. For example, there is solid evidence linking IL-2 receptor subunit alpha (IL-2ra) increasing variants to elevated risk for multiple sclerosis (MS) and Crohn’s disease (CD).^17^ Aldesleukin, a recombinant form of IL-2 approved for cancer indications, is currently under investigation in a phase-2 clinical trial for CD (ClinicalTrials.gov ID: NCT04263831).^43^ Also, compounds targeting IL-1 signaling, anakinra or canakinumab, represent established treatment algorithms for inflammatory joint diseases like rheumatoid arthritis (RA) or juvenile arthritis.^44,45^

**Figure 8.**
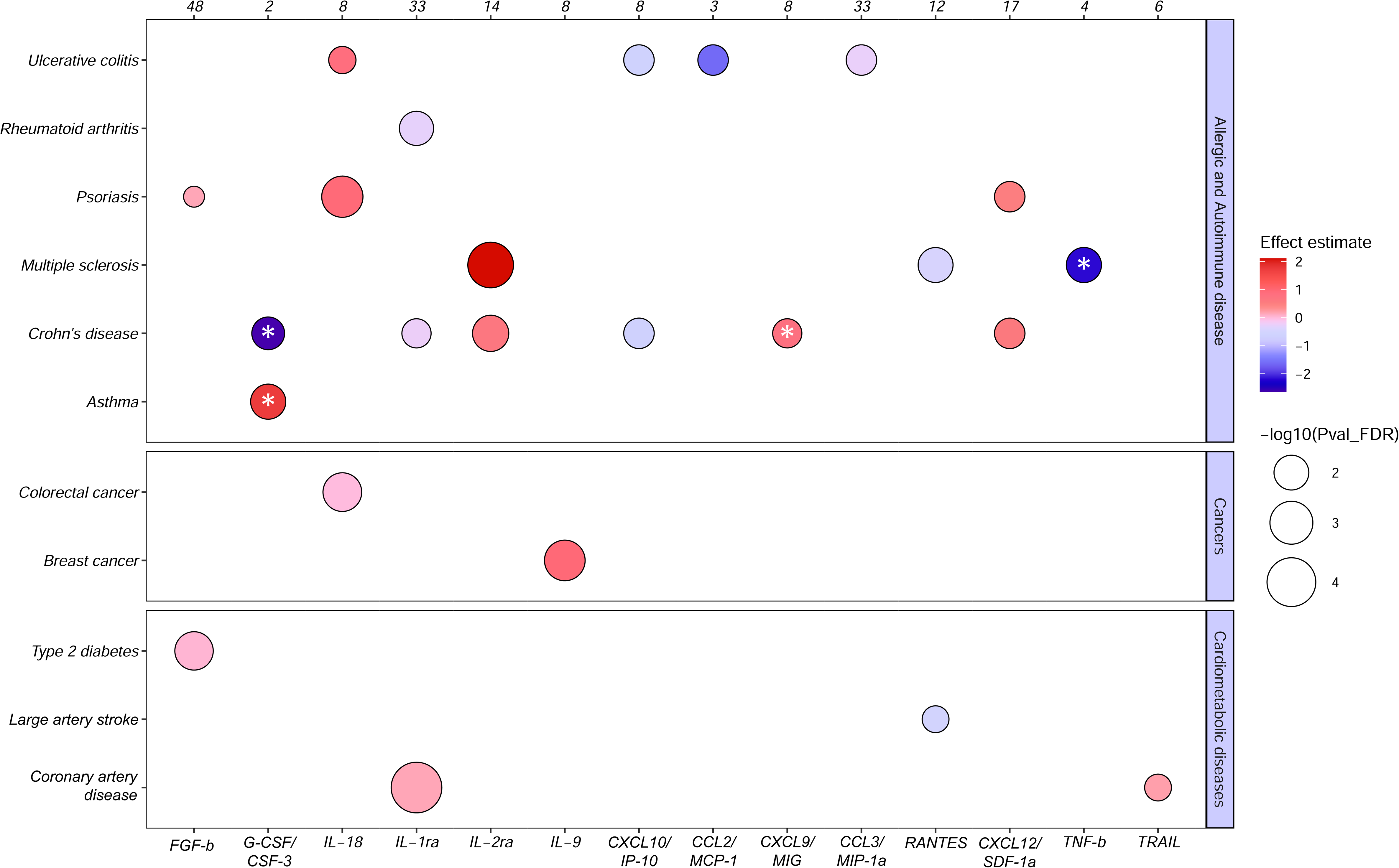
*Cis*-Mendelian randomization associations and colocalization analyses between genetically proxied cytokine levels and disease risk. Significant associations between circulating cytokine levels and disease outcomes are shown for allergic and autoimmune, cardiometabolic, and cancer outcomes. Effect sizes and log-transformed, Benjamin-Hochberg corrected p-values are illustrated by color gradient and circle size, respectively. Only cytokines and disease endpoints with at least 1 significant association are depicted. Numbers at the top indicates average number of *cis*-acting genetic variants used as instruments in MR analyses. Stars highlight significant genetic colocalizations (posterior probability of association >80%) for shared causal variants between circulating cytokine levels and disease risk.

Of the 24 signals, 4 also showed evidence of significant colocalization, that is a PP of association >80% for shared causal variants between cytokine levels and disease outcomes (**Figure 8**, **Supplementary Table S13**), thus providing even stronger evidence for causality. These included associations of higher genetically proxied G-CSF/CSF-3 levels with asthma, lower genetically proxied G-CSF/CSF-3 and higher genetically proxied CXCL9/MIG levels with CD, as well as lower genetically proxied TNF-b levels with MS. Furthermore, the association between genetically proxied IL-1 receptor antagonist (IL-1ra) levels and lower risk of RA reached a PPA of 68% for a shared causal variant in colocalization analysis. These results are consistent with data from preclinical studies,^46–48^ observational studies in humans,^49–53^ and clinical trials,^54,55^ thus providing support for potentially promising targeted immunotherapies for these indications.

### Integration of cytokine-disease MR and TWAS-MR results implicates additional mediators of disease mechanisms that could represent promising drug targets

As a last step, we aimed to integrate the cytokine-disease MR results with the TWAS MR results with the goal of also detecting upstream regulators of the potentially causal cytokines. We performed MR analyses between genetically proxied expression of genes significantly associated with G-CSF/CSF-3, CXCL9/MIG, and TNF-b in our TWAS-MR analyses and the associated disease outcomes. We found that higher genetically proxied expression of *PPP1R37* is associated with lower levels of G-CSF/CSF-3, as well as with a lower risk of asthma **(Figure 9a)**. We also found higher genetically proxied expression of *TRAFD1* to be associated with higher CXCL9/MIG levels and higher risk of CD **(Figure 9b)**.

**Figure 9.**
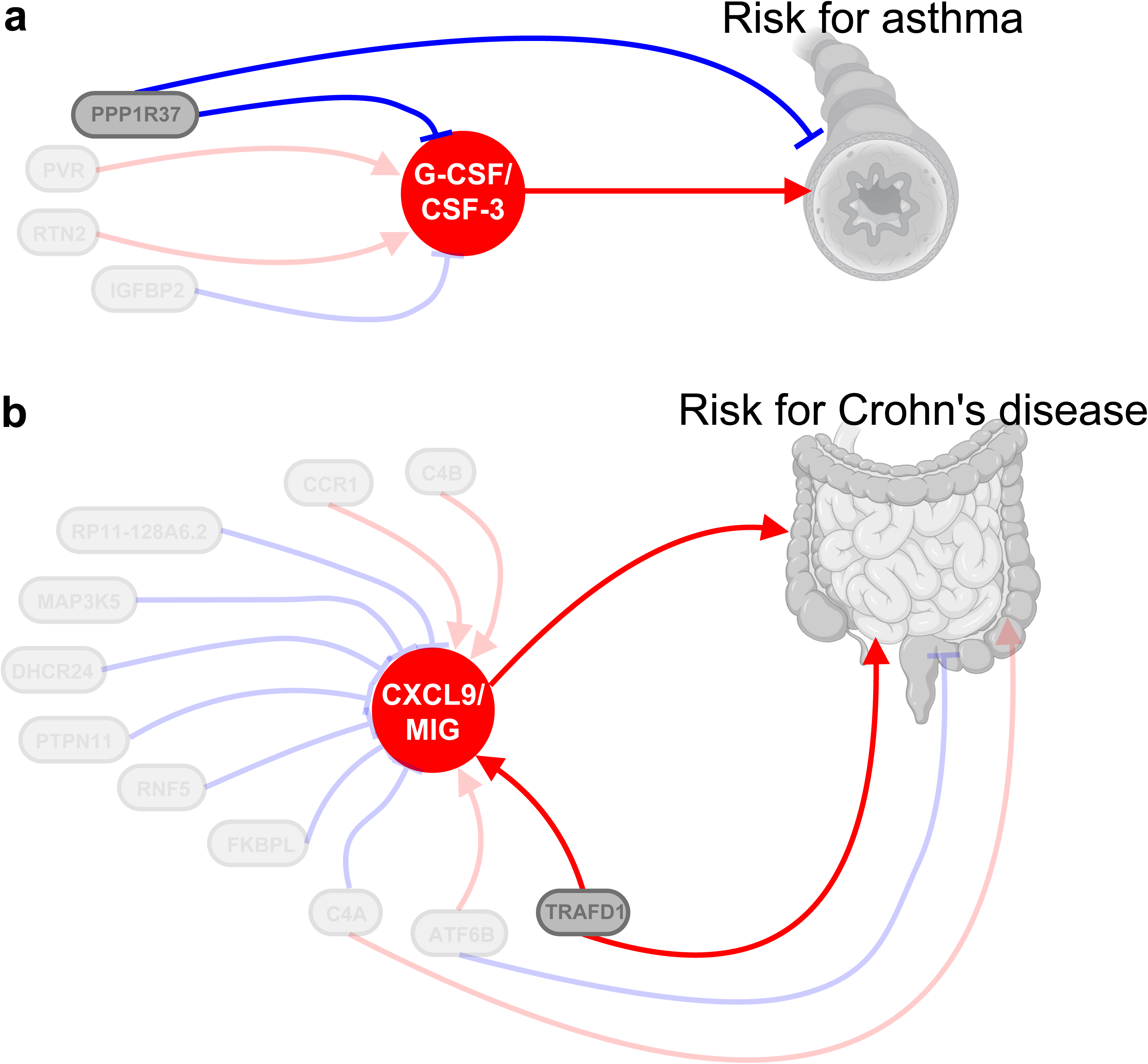
Causal associations between genetic regulators for cytokines, circulating cytokine levels and disease risk. (**a**) Genetically proxied mRNA for *PPP1R37*, *PVR*, *RTN2* and *IGFBP2* affect serum G-CSF/CSF-3 levels leading to increased risk for asthma. In turn, *PPP1R37* directly lowers disease risk for asthma. (**b**) Genetically proxied mRNA for 11 genes underlying CXCL9/MIG serum levels differentially affect circulating cytokine concentrations which influence the risk for Crohn’s disease. Independently, *TRAFD1*, *ATF6B* and *C4A* also modulate disease risk for Crohn’s disease.

## Discussion

Pooling data from up to 74,783 patients from 3 independent GWAS cohorts, we identified 169 independent genomic loci influencing the circulating concentration of 40 cytokines, 156 of which have not been associated with circulating cytokine levels in previous GWASs. Integrating our results with transcriptomic data, a TWAS-MR analysis revealed 245 potentially causal associations between gene expression of mostly immunoregulatory genes in peripheral blood and circulating cytokine levels. Analyzing regulatory interactions between cytokines, we found TNF-b, VEGF, and IL-1ra as master controllers of the circulating levels of multiple cytokines. Finally, we provide genetic evidence that the circulating levels of 3 cytokines (G-CSF/CSF-3, CXCL9/MIG, TNF-b) might be causally involved in the pathogenesis of asthma, CD, and MS, thus offering insights for the development of more specific immunotherapies.

Our MR and colocalization analyses provided evidence for potential causality for four cytokine-indication pairs, thus offering genetic support for potentially promising targeted immunotherapies for asthma, CD, and MS. These results are highly consistent with preclinical, epidemiological, and occasionally clinical data. For example, G-CSF/CSF-3 is a pro-inflammatory cytokine involved in neutrophil differentiation and systemic mobilization and has been implicated in the pathogenesis of neutrophilic atopic asthma.^46,56^ Several preclinical studies in asthma models showed that blockage of upstream inductors or the receptor of G-CSF/CSF-3 reduced circulating cytokine levels, alleviated the airway inflammatory response, and improved disease outcome.^46–48,57^ Furthermore, G-CSF/CSF-3 levels in the sputum of asthma patients have been suggested as a marker of airway neutrophilic inflammation.^58^ Our results provide genetic support for the concept of targeting G-CSF/CSF-3 in asthma, potentially focusing on patients with neutrophilic asthma.

Using integrated results from our GWAS and TWAS findings, we identified an upstream mechanism of yet unknown relevance: higher *PPP1R37* gene expression was associated with reduced G-CSF/CSF-3 serum levels and lower risk of asthma. The gene encodes a regulatory subunit that acts as phosphatase inhibitor and has, so far, not been associated with airway diseases.^59^ Prveious studies investigating related regulatory subunits have unveiled potential biological mechanisms through which these subunits may influence the immune response^60,61^ and genetic studies have provided support for the significance of other protein phosphatase regulatory subunits as contributing factor to airway diseases.^62–64^ For example, genetically proxied expression of *PPP1R**3D*** was associated with disease characteristics of asthma including mucosal immunity, cell metabolism, and airway remodeling and predicted responsiveness to omalizumab therapy.^62^ Although the specific biological mechanism underlying our finding is unknown, one might speculate that the diverse range of functions associated with PP1, including cell progression, apoptosis, and muscle contraction, might underlie the observed findings.

The MR results supported a connection between genetically proxied circulating levels of 6 cytokines and CD. Among them, associations of higher CXCL9/MIG and lower G-CSF/CSF-3 with CD were also supported by colocalization evidence. CXCL9/MIG, a proinflammatory IFNg-induced CXC-chemokine, is released by various immune cells including macrophages to attract and activate T-cells and forms together with neighboring CXCL10/IP-10, CXCL11/IP-9 and their cognate CXCR3 receptor an axis with specific relevance in inflammatory bowel disease.^65,66^ In clinical studies, elevated CXCL9/MIG serum levels have been associated with CD relapses, affirming CXCL9/MIG as a risk factor in CD.^53^ Apart from pro-inflammatory actions, exogenous administration of G-CSF/CSF-3 has been associated with immunoregulatory effects, such as modulation of T-cell responses.^67^ Two open-label studies have indeed demonstrated that subcutaneous G-CSF/CSF-3 is effective in inducing clinical remission, fostering mucosal healing, and normalizing cell counts and cytokine responses in CD patients.^54,55^

Integrating our results linking CD with circulating CXCL9/MIG levels, we identified genetically proxied expression of *TRAFD1* as a potential upstream causal regulator of CXCL9/MIG levels and subsequently risk of CD.^53,68,69^ TRAFD1 binds as homodimer or in interaction with TRAFD2, to TNF receptors, impacting pro-inflammatory cytokine production and modulating inflammatory responses in immune cells.^32,33,70^ In celiac disease, *TRAFD1* was recognized as an upstream regulator of IFNg signaling and thereby activating cytotoxic T-cells, an important pathomechanism.^34^ The identified function of *TRAFD1* as inductor of IFNg signaling aligns well with the literature and also with our findings showing increased CXCL9/MIG serum levels underlying *TRAFD1* expression.^66^ Substantiating the risk-increasing associations in our analysis, elevated expression of *TRAFD1* and *TRAFD2* was noticed in acutely inflamed mucosal biopsies of CD patients.^71^ Together these reports confirm our findings for the importance of *TRAFD1* in the pathology of CD and as modulator of inflammatory reactions through effects on cytokine levels.

Our analysis also provided evidence for an inverse association between genetically proxied circulating TNF-b and MS risk. TNF-b, is a pro-inflammatory cytokine within the TNF superfamily with substantial genetic correlation to TNF-a and binding affinity to pro-inflammatory as well as anti-inflammatory TNF receptors.^72^ In a randomized phase 2 trial the TNF-inhibitor lenercept was tested for safety and efficacy in MS but had to be terminated prematurely after the interim analysis detected a dose-dependent increase in the frequency and severity of MS exacerbations.^73^ In contrast to the TNF blockers infliximab, adalimumab and golimumab, lenercept provides equal inhibitory efficacy for TNF-a and TNF-b.^72,74^ While TNF inhibition has demonstrated success in treating autoimmune diseases such as rheumatoid arthritis or psoriasis, patients undergoing anti-TNF therapy for these indications are at risk for developing demyelinating CNS lesions, indicating a disease-specific effect.^40,75^ Supporting the clinical findings, GWAS studies in MS identified TNF lowering alleles for both cytokines TNF-a and TNF-b that were associated with higher risk for MS.^23,76^ We provide additional genetic evidence in line with observational, clinical, and GWAS findings for a potentially protective role of TNF-b in MS.

In our network analysis, over 80 percent of the significant cytokine-cytokine interactions led to an increase in downstream cytokine concentrations. Given that the majority of the involved cytokines were pro-inflammatory implies a self-perpetuating feedback mechanism leading to strong inflammatory responses and suggests a global trend where cytokines mutually activate each other to amplify their immune reaction. For the IL-1ra and TNF-b network, specifically, we successfully expanded the list of downstream affected cytokines. TNF-b exhibited notable downstream effects on 13 other cytokines, signifying its role as a master regulator. This observation aligns with existing reports on *TRAFD1’s* downstream effects, influencing various cytokine-encoding genes like CXCL10/IP-10 and IFNg.^34^

Our study has limitations. First, our meta-analysis was based on 3 cohorts that used different affinity-based assaying approaches for quantifying circulating cytokines. The different approaches might yield varying measurements for the same proteins with only moderate correlations across the assays.^18^ This might explain the difference in replication rates across the 3 cohorts. Interestingly, we found a higher replication rate for signals detected with the Olink assay. In a previous cross-assay comparison between Olink and SomaScan, the proportion of assays with detected pQTLs was also higher with the Olink-based assay.^18^ The differences across the panels should be further explored at a larger scale to explore the extent to which it would be possible to scale genetic explorations across cohorts utilizing different proteomic platforms. Second, due to differences in reporting of effect sizes for genetic variants across the GWAS source data, we could perform only p-value-based meta-analyses and only indirectly estimate the pooled effect sizes based on the derived p-values and the variant allele frequencies. Inaccuracies in this estimation could influence downstream analyses heavily relying on effect sizes. Third, due to data availability, our analyses were based on 40 selected cytokines. Future endeavors utilizing solely high-throughput proteomic data could scale up to analyses including more inflammatory proteins. Fourth, our analyses are based on individuals of European, Finnish, and Icelandic ancestry and as such might not be generalizable to individuals of a different ancestry background. Fifth, due to the large number of cytokines we adjusted the significance level for multiple testing which might have neglected important findings due to non-significance. Using a hypothesis-driven approach, future studies should follow-up on our results to identify additional targets we might have missed for a comprehensive view around our findings. Sixth our GWAS meta-analysis was based on population-based cohorts without predominant inflammatory diseases. Genetic variants might influence cytokine levels in specific contexts such as a response to infection or other pro-inflammatory stimuli; our approach could not detect such signals.

## Conclusion

In conclusion, our study, leveraging data from 74,783 individuals across 3 cohorts, identified 169, mostly novel, genomic loci influencing circulating cytokine levels. Follow-up analyses of the detected signals reveal interesting underlying pathways, which enhance our understanding of the biology of the immune response. Integrating our data with genetic data for human disease risk, our analyses suggest potential targets like G-CSF/CSF-3, CXCL9/MIG, and TNF-b for immune-related diseases including asthma, Crohn’s disease, and multiple sclerosis, warranting further exploration in clinical trials. The summary statistics from our study offer a valuable resource for future omics analyses, aiding data integration for the identification of potential drug targets for human diseases.

## Acknowledgements

This work is supported by the German Research Foundation (Deutsche Forschungsgemeinschaft, DFG) by an Emmy Noether grant (GZ: GE 3461/2-1, ID 512461526 to MG), by a clinician-scientist grant to MG and project grants to JB and MD from the Munich Cluster for Systems Neurology (EXC 2145 SyNergy, ID 390857198), and by CRC1123 project A3 to JB. Additional support comes from a research grant from the Fritz-Thyssen Foundation (Ref. 10.22.2.024MN to MG) and a research fellowship by the Hertie Foundation (Hertie Network of Excellence in Clinical Neuroscience, ID P1230035 to MG).

## Materials & Methods

### Study populations and design

The study cohorts and a flowchart of the study design are depicted in **Figure 1**. We downloaded publicly available GWAS summary statistics for the circulating levels of up to 40 cytokines from 3 independent cohorts. Details of the study protocols have been published elsewhere.^17,77,78^ Human genome assembly GRCh37 (hg19) from Genome Reference Consortium was used for genomic positioning.^79^ Before further computations, all 3 databases were harmonized regarding data structure. For the GWAS meta-analyses and downstream computations we included all cytokines that were available in at least 2 cohorts. To ensure that the available cytokines were identical between cohorts we used information provided on the NIH (https://www.ncbi.nlm.nih.gov/gap/) and GeneCards (https://www.genecards.org/) websites and verified synonyms and aliases in the abbreviated and full names of the cytokines. We excluded 1 cytokine (IL-12) from the analyses, because different subunits of the protein were quantified in the two cohorts (IL-12p70 in YFS & FINRISK and IL-12p40 in deCODE).

### YFS and FINRISK

Genomic data for 40 cytokines were drawn from up to 8,293 individuals of Finnish background that were included in the YFS & FINRISK cohorts 1997 and 2002, respectively.^17^ The mean age across all studies was 49 years (standard deviation 8 years). The cytokine measurements were carried out in EDTA plasma for the FINRISK 1997 cohort, in heparin plasma for the FINRISK 2002 cohort and in serum for the YFS cohort using cytokine Luminex®L-based multiplex immunoassays from Bio-Rad®L. Genotyping was completed using the Illumina HT12 platform for the YFS study and the Illumina 670k HumanHap array for both FINRISK studies. Imputation was performed using the 1000 Genomes reference panel across all cohorts.^79^ The GWAS meta-analyzing all 3 studies normalized the cytokine distribution using inverse transformation and adjusted the genetic analyses for age, sex and ancestral principal components 1-10. The reported effect sizes were scaled per standard deviation increment in inverse-transformed cytokine levels.

### SCALLOP Consortium

Genomic data for 16 cytokines were drawn from up to 30,931 individuals with European background from the SCALLOP consortium, a collaborative framework analyzing gene-protein associations across 13 studies.^78^ The cytokine measurements were carried out in plasma samples using the proximity extension assay-based Olink^®^ platform. Genotyping methods across the studies included Cardiometabochip, Immunochip, PsychChip, Illumina HumanCoreExome, Illumina OmniExpress, Metabochip, Illumina OmniExpress 2.5, Affymetrix Axiom UK Biobank array, HumanCytoSNP-12 BeadChip, HapMap300v2, Human Exome, Illumina HumanOmniExpressExome-8 v1, Illumina HumanHap300v1, Omni1, OmniX, Illumina HumanHap300v1 and Infinium PsychArray-24 v1.2. Imputation was performed using the following panels: 1000G phase v5, 1000G phase v3, UK10K reference panel, HRC, HRC r1.1. The GWAS meta-analyzing all 13 studies adjusted the cytokines for age, gender, site, OLINK batch, Olink plate, MDS components, storage time, bleed to processing time (days), smoking status, oral contraceptive usage, blood cell counts, season of venipuncture and ancestral principal components 1-10. The log2-based normalized expression values (NPX) for each protein were rank-based inverse normal transformed and standardized to units of standard deviation.

### deCODE

Genomic and proteomic data for 39 cytokines were taken from 35,559 Icelandic individuals included in deCODE.^77^ The mean age was 55 years (standard deviation 18 years). The cytokine measurements were carried out in plasma samples using the aptamer-based SOMAScan^®^ assay. Genotyping was completed using Illumina SNP Chip. Imputation was based on an in-house developed whole genome sequencing reference panel. The genetic analyses were adjusted for age and sex. Cytokine measurements were normalized using rank-inverse normal transformation and standardized to standard deviation increment. To allow alignment with other datasets, we excluded all SNPs that were not covered by the 1000 Genomes reference panel.

### Cross-assay comparisons in the genetic architecture of circulating cytokines

To explore differences in the genomic architecture of cytokines levels between the 3 studies that applied different measurement assays, we compared the proportion of overlapping SNPs between datasets confined to significant (p-value < 0.05) and directionally concordant (same direction of effect estimates across all 3 databases) variants. Using the raw data, we analyzed overlapping SNPs by taking one dataset as reference comparing it to the other two.

### GWAS meta-analyses

We performed fixed-effects inverse variance-weighted meta-analysis for each cytokine across the available cohorts using METAL software (v.2011-03-25, number of cohorts and sample sizes per cytokine GWAS are provided in **Supplementary Table S14**).^80^ Due to differences in scaling of the derived effect estimates across the 3 datasets, we applied a z-score-based meta-analysis (SCHEME SAMPLESIZE). Subsequently, we estimated standardized beta coefficients using p-values, minimum allele frequency, and direction of effects, weighted according to sample sizes, as previously described.^81^ For estimation of heterogeneity of effect sizes between data sources we calculated chi-square test statistics for all included markers. To control for genomic inflation we calculated lambda statistics for each cytokine (**Supplementary Table S3**).^82^ Significant variants were defined based on the established genome-wide significance level (p<5×10^-8^). To detect independent variants following correction for linkage disequilibrium, we clumped across the significant ones using clump_data (TwoSampleMR R package version 0.5.6) at an r^2^<0.001 based on the European 1000 Genomes Project reference panel.^79^ We defined independent loci as SNPs that were separated by more than 1 Mb from the next SNPs in the 3’ and 5’ direction, as reported earlier.^83^

### Linkage disequilibrium score regression (LDSC)

Using the LD score v1.0.1. tool we applied LDSC regression with reference data from the European 1000 Genomes project for calculation of cross-trait LDSC genetic correlations between all 40 cytokines using the meta-analysis results.^79,84–86^

### Fine mapping, functional annotation, pathway and gene-set analysis

To identify causal variants responsible for variations in circulating cytokine concentrations, we investigated significant loci associated with cytokines. We employed PLINK v1.9 to compute LD score correlation matrices and further refined the results using SuSiE (susieR R package version 0.12.16) to derive sets of variants, ensuring the inclusion of at least one causal variant with a cumulative probability ≥95%.^87,88^ Subsequently, the causal variants were utilized to estimate the total variance explained by the identified loci for individual cytokines.^89^ For functional analyses we used phenoscanner (MendelianRandomization R package version 0.6.0) which ascribes functional consequences (intron, intergenic, exon, upstream, downsteam, etc.) of single variants using positional mapping (physical distance).^90,91^ Gene-property analyses was conducted for identification of the tissue specificity of cytokines using the FUMA Gene2Func web database.^92^ Lastly, MAGMA gene-based and gene-set analyses were conducted. Gene-based analysis initially calculates p-value association tests for variants mapped to protein coding genes which are then used to calculate gene-set p-values in the gene-set analysis. Using predefined gene-sets, variants with significant associations to genes can then be analyzed to determine their underlying functional or process-related feature, i.e. gene-sets belonging to molecular functions or biological processes.^93^

### Transcriptome-wide Mendelian randomization analysis (TWAS-MR)

To further explore whether variant effects on expression of specific genes underlie the genetic underpinnings of circulating cytokine levels, we performed transcriptome-wide inverse variance-weighted 2-sample MR analysis, as has been previously described.^94^ Sensitivity analyses were conducted using MR Egger regression and the weighted median estimator to control for horizontal pleiotropy.^95,96^ For calculation of effect estimates we used the mr command from the TwoSampleMR R Package (TwoSampleMR version 0.5.6) with *cis*-expression quantitative trait loci (eQTL) gene instruments from the eQTLGen Consortium as exposure (clumped at r^2^<0.01) and the GWAS meta-analysis results of our cytokine panel as outcome. The eQTL consortium included 31,684 individuals of primarily European ancestry (detailed methods have been described previously and are available online).^22^

### Mendelian randomization analyses

We performed MR analyses exploring (i) the effects of circulating cytokine levels on other cytokines, (ii) the effects of circulating cytokine levels on allergic and autoimmune, cardiometabolic, and cancer disease endpoints, (iii) and, depending on the outcomes of the 2^nd^ MR analyses the effects of gene transcripts (eQTL) upstream of promising cytokines on allergic and autoimmune endpoints. We used *cis*-acting variants as genetic instruments for our MR analyses, as they are associated with a lower risk of pleiotropic effects when compared to *trans*-acting variants.^97^ We filtered the GWAS meta-analysis results for variants within 300 kb around the gene encoding the respective cytokine. We selected variants associated at p<5×10^-5^ and clumped the data at r^2^<0.1. We applied fixed-effects inverse variance-weighted MR analysis as our main analytical approach.^94^ Again, MR egger regression and the weighted median estimator were used as sensitivity analyses.^95,96^ After harmonization of the effect alleles across cytokines we used mr command from the TwoSampleMR R Package (TwoSampleMR version 0.5.6) to extract the respective effect estimates.

### Disease outcome GWASs

For the disease endpoints, we downloaded summary level data from the largest publicly available GWAS and performed MR analyses for 3 independent disease groups. For allergic and autoimmune phenotypes we analyzed asthma (121,940 cases, 1,254,131 controls)^98^, Crohn’s disease (5,956 cases, 14,927 controls)^99^, ulcerative colitis (6,968 cases, 20,464 controls)^99^, multiple sclerosis (47,429 cases, 68,374 controls)^100^, psoriasis (4,815 cases, 415,646 controls)^101^, and rheumatoid arthritis (14,361 cases, 43,923 controls)^102^. For cardiometabolic phenotypes we analyzed peripheral vascular disease (31,307 cases, 211,753 controls)^103^, coronary artery disease (60,801 cases, 123,504 controls)^104^, large artery stroke (9,219 cases, 1,503,898 controls)^105^ and diabetes mellitus type II (242,283 cases, 1,569,730 controls)^83^. For cancer phenotypes we analyzed breast cancer (133,384 cases, 113,789 controls)^106^, colorectal cancer (5,657 cases, 372,016 controls)^107^, lung cancer (29,266 cases, 56,450 controls)^108^, non-Hodgins lymphoma (2,400 cases, 410,350 controls)^109^, and skin cancer (23,694 cases, 372,016 controls)^107^. The data sources are detailed in **Supplementary Table S15**.

### Colocalization analysis

To analyze shared causal variants between SNPs for circulating cytokines and disease outcomes showing significant associations in MR analyses, we used the “coloc” v3 R package. COLOC is a variant colocalization method that performs tests on shared causal variants in the locus. Colocalization methods consider the GWAS and disease outcome summary statistics at a locus jointly and probabilistically test if the two signals are likely to be generated by the same causal variant. ^110^ We used the meta-analyses summary statistics for the significant cytokines restricted to a flanking region ±300Kb around the genetic location of each cytokine and mapped disease-associated variants by their rsID.

### Database search

To assess previously reported associations a database search was conducted using the NHGRI-GWAS catalogue^111^ on February 15^th^, 2023. We analyzed our GWAS hits for assocations with any of the 40 cytokines reported here **(supplementary Table S4),** restricting the results for European-ancestry associations.

## Data Availability

GWAS meta-analysis summary statistics will be available at GWAS catalog

## Data availability

The data sources used in the current study are publicly available (download links for the summary statistics are available in **supplementary Table S14**). Ethical approval was not required due to usage of publicly available summary-level data.^17,77,78^ GWAS meta-analysis summary statistics will be available at GWAS catalog.

## Code availability

Codes used to generate the results and figures are available on request from the corresponding author.

